# Retrospective Exploratory Analysis of Task-Specific Effects on Brain Activity after Stroke

**DOI:** 10.1101/2021.08.20.21260371

**Authors:** Marika Demers, Rini Varghese, Carolee J. Winstein

**Affiliations:** Division of Biokinesiology and Physical Therapy, Herman Ostrow School of Dentistry, University of Southern California, 1540 Alcazar St, Los Angeles, (CA), USA, 90089; Department of Neurology, Keck School of Medicine, University of Southern California, 1975 Zonal Ave, Los Angeles (CA), USA, 90033

**Keywords:** neurorehabilitation, neuronal plasticity, upper extremity, stroke, motor cortex, magnetic resonance imaging

## Abstract

**Background:** Evidence supports cortical reorganization in sensorimotor areas induced by constraint-induced movement therapy (CIMT). However, only a few studies examined the neural plastic changes as a function of task specificity. This retrospective exploratory analysis aims to evaluate the functional brain activation changes during a precision and a power grasp task in chronic stroke survivors who received two-weeks of CIMT compared to a no-treatment control group.

**Methods:** Fourteen chronic stroke survivors, randomized to CIMT (n=8) or non-CIMT (n=6), underwent functional MRI (fMRI) before and after a two-week period. Two behavioral measures, the 6-item Wolf Motor Function Test (WMFT-6) and the Motor Activity Log (MAL), and fMRI brain scans were collected before and after a two-week period. During scan runs, participants performed two different grasp tasks (precision, power). Pre to post changes in laterality index (LI) were compared by group and task for two predetermined motor regions of interest: dorsal premotor cortex (PMd) and primary motor cortex (MI).

**Results:** The CIMT group showed significant improvements in both the WMFT-6 and the MAL post-intervention, whereas the non-CIMT group showed improvements in only the MAL. Two weeks of CIMT resulted in a relative increase in activity in a key region of the motor network, PMd of the lesioned hemisphere, under precision grasp task conditions compared to the non-treatment control group. No changes in LI were observed in MI for either task or group. Conclusions: These findings provide evidence for task-specific effects of CIMT in the promotion of recovery-supportive cortical reorganization in chronic stroke survivors.

## 1. Introduction

Growing evidence suggests that rehabilitation interventions that harness motor practice can drive the brain’s restorative capacity. In a seminal study, Nudo et al.(1996) demonstrated evidence for cortical reorganization in the areas representing the distal forelimb in primates, not simply from spontaneous recovery, but as a result of motor skill training. Similar training-induced improvements in motor function and positive restorative neural plasticity in spared cortical regions have also been observed in other animal models (Castro-Alamancos and Borrell, 1995; Jones *et al*., 1999; Biernaskie and Corbett, 2001).

Intense task-specific motor training, such as Constraint-Induced Movement Therapy (CIMT), has been shown to reduce motor impairments in the paretic arm (Wolf *et al*., 2008; Corbetta *et al*., 2015; Kwakkel et al., 2015) and is thought to mediate sensorimotor recovery through experience-dependent cortical reorganization (Schaechter et al., 2002; Wittenberg *et al*., 2003; Liepert *et al*., 2006; Laible *et al*., 2012). Although CIMT, by definition, consists of a mitt constraint applied to the less-impaired arm, its most effective ingredient may be the *intensity* of practice (∼60 hours over 2 weeks) of increasingly difficult tasks, known as “shaping”, combined with a transfer package for at-home practice (Winstein *et al*., 2003; Kaplon *et al*., 2007; Taub *et al*., 2013; Winstein and Wolf, 2014).

Intensive shaping practice can induce experience-dependent plasticity in the primary and secondary motor cortical regions of the lesioned hemisphere (Johansen-Berg et al., 2002; Wittenberg *et al*., 2003; Kwakkel *et al*., 2015). In the last decade, there is evidence that the behavioral improvements induced by CIMT are associated with cortical reorganization in sensorimotor areas (Levy *et al*., 2001; Johansen-Berg *et al*., 2002; Jang *et al*., 2003; Wittenberg *et al*., 2003; Kim *et al*., 2004; Schaechter and Perdue, 2008; Laible *et al*., 2012). For example, a study using transcranial magnetic stimulation showed increased motor map size in the cortex of the lesioned hemisphere in stroke survivors undergoing CIMT compared to a control group (Wittenberg *et al*., 2003).

One of the key tasks practiced during CIMT is the precision grasp, where the object is pinched between the flexor aspect of the fingers and that of the opposing thumb (Napier, 1956). Precision grasp involves finger individuation, and anticipatory movement planning to perform goal-directed, skilled actions. In contrast, power grasp involves undifferentiated finger and thumb movements of the whole hand for force control (Napier, 1956). Two cortical brain regions associated with precision and power tasks are the dorsal premotor cortex (PMd) and the primary motor cortex (MI), respectively. PMd is active during movement preparation, action selection and online control of reaching movements (Kantak *et al*., 2012). MI is associated with force control and movement execution (Cramer *et al*., 2002; Johansen-Berg *et al*., 2002). Our previous work demonstrated that a 2-week CIMT intervention resulted in improved anticipatory planning of hand posture selection, particularly in situations that require precision grasp actions, and improved movement time in reach-to-grasp tasks (Tan *et al*., 2012). We reason that dexterous and manipulative tasks practiced in the context of CIMT would more likely engage circuits involved in anticipatory planning than primarily force control or strength tasks (Muir and Lemon, 1983; Carey, Bhatt and Nagpal, 2005; Kantak *et al*., 2012).

Functional brain imaging studies of stroke motor recovery have primarily used undifferentiated finger tasks. Only a few studies have examined neural plastic changes as a function of task specificity and motor learning (Carey, Bhatt and Nagpal, 2005; Schaechter and Perdue, 2008). This provoked us to reanalyze an unpublished dataset from a companion study of the EXCITE trial (Winstein *et al*., 2003) in pursuit of evidence for cortical reorganization induced through task-specific training in chronic stroke survivors. We selected two different grasp tasks: a precision grasp task involving force modulation through differentiated finger movement and a power grasp task involving force modulation through undifferentiated finger movement. A comparison of the neural activation pattern elicited from these two fundamentally different grasp tasks would allow a direct examination of the task-specific effects of CIMT and provide evidence about how task-specific training might modulate recovery-supportive functional plasticity.

This proof-of-concept exploratory analysis aims to evaluate the functional brain activation changes during a precision and a power grasp task in chronic stroke survivors who received two weeks of CIMT compared to a no-treatment control group. We selected two motor regions of interest (ROIs), PMd and MI, for their significant involvement in motor network changes obtained from both preclinical animal model and human clinical reports of upper limb recovery after stroke (Nudo, 2007; Buma *et al*., 2010; Calautti *et al*., 2010). We hypothesize that the precision task, more than the power task, would elicit greater brain activation of PMd of the lesioned hemisphere in the CIMT compared to the non-CIMT group. We expect differential group effects of task for change in activation laterality index for both ROIs.

## 2. Material and methods

### 2.1 Participants

Fourteen chronic stroke survivors (5-12 months post-stroke) with mild-moderate motor impairments were randomized to CIMT (n=8) or non-CIMT (n=6). Participants with functional magnetic resonance imaging (fMRI) safety contraindications and severe cognitive impairments were excluded. Only participants with a nearly complete set of evaluable behavioral and fMRI for baseline and immediate post-intervention visits were included in this retrospective analysis. All participants signed an informed consent approved by the Health Sciences Institutional Review Board of the University of Southern California.

### 2.2 Intervention

The CIMT group completed the signature CIMT protocol, which consisted of a mitt constraint applied to the less-impaired arm and intensive, supervised task-specific training 6 hours/day, 5 days/week, for 2 weeks (Winstein *et al*., 2003). The non-CIMT group completed the behavioral testing and fMRI scans two weeks apart, but received no formal rehabilitation training.

### 2.3 Behavioral measures

Before and after a two-week period, the 6-item Wolf Motor Function Test (WMFT-6) and the Motor Activity Log (MAL) were administered by a blinded assessor. The WMFT-6 is a subset of the WMFT, includes six time-based precision and dexterous tasks and is strongly correlated with the 15-item WMFT (r=0.61, p=0.02; Wolf *et al*., 2005). The MAL, a structured interview is a self-reported measure in which participants rate the quantity, Amount of Use (AOU) and Quality of Movement (QOM) of the paretic arm during 15 everyday activities performed over the past 3 days (Taub *et al*., 1993; Uswatte and Taub, 1999). Each activity is scored on a 6-point scale with higher scores indicating better performance. The upper limb Fugl-Meyer motor assessment (FMA; Fugl-Meyer *et al*., 1975) was also administered at baseline to determine impairment level (0: severe motor impairments to 66: normal function).

### 2.4 fMRI task probe description

Participants repetitively compressed either a vertically mounted plastic tube with the index and middle fingers against the thumb in a precision grasp posture (precision grasp), or a vertically mounted rubber bulb with all digits in a power grasp posture (power grasp), (Fig. 1A, B, D). A custom-built fMRI-compatible apparatus with the two grasp units was connected to a pneumatic pressure transducer. The output from the pressure transducers were collected electronically using custom MATLAB software for subsequent off-line analysis (dataWizard, v. 0.9; https://sites.google.com/site/ucladatawizard/). Prior to each fMRI session, participants practiced each grasp task with the paretic hand, in the supine position, outside of the scanner to: (1) establish and ensure across-session consistency of force level and rate; (2) reduce mirror movements of the opposite arm and associated head, elbow and shoulder movements. During the practice period, participants were offered visual feedback about force production to maintain a consistent pre-specified force and rate (Fig. 1C). Once a participant was able to perform each grasp task with a consistent pressure and rate without feedback, they were scheduled for the fMRI sessions. During the scanning, participants were reminded to maintain the same pressure and rate that they had practiced—i.e., 50% of their maximum pressure and 75% of their maximum rate, without feedback. These levels were chosen to 1) account for a range of individual differences in force capability and motor control and 2) avoid fatigue during the fMRI scanning blocks.

**Figure 1.**
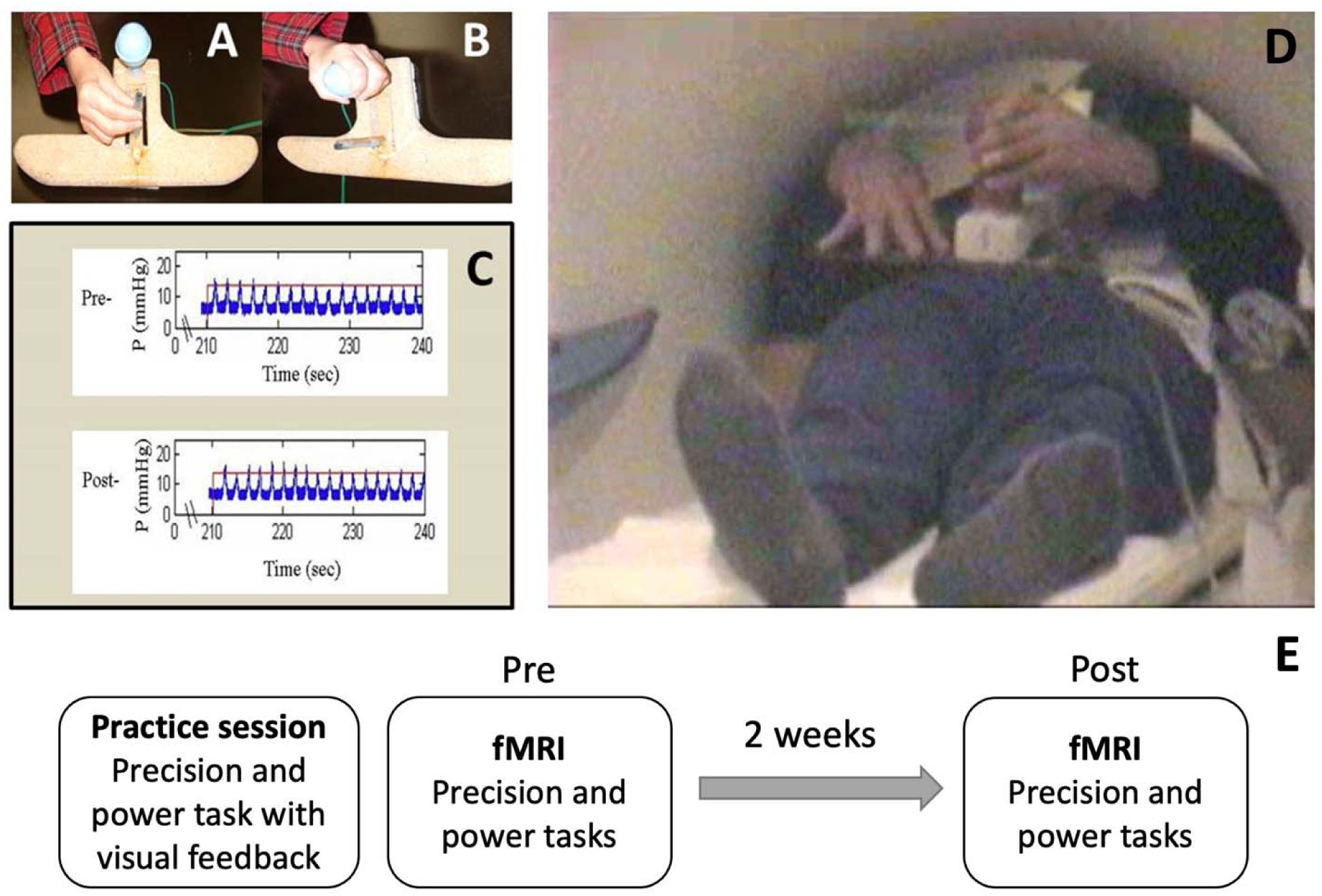
Motor Activation Tasks performed in the functional magnetic resonance imaging (fMRI) scanner. Task apparatus MRI safe device with two pneumatic pressure sensors for (A) precision grip and (B) power grasp. C. Sample pre- and post-pressure and rate graphs demonstrating participant ability to maintain 50% of their predetermined maximum pressure and 75% of their predetermined maximum rate throughout each fMRI session. D. Participants were positioned to minimize movement within the scanner. No visual feedback was offered during the scan. E. Study timeline.

### 2.5 fMRI data acquisition

Functional and structural images were acquired using a 1.5 Tesla Siemens Sonata scanner at the Brain Mapping Center, University of California, at Los Angeles. Auto-shimming was conducted at the beginning of the scan to correct magnetic inhomogeneity. All functional and structural images covering the whole brain were acquired parallel to the anterior-posterior commissure line using a sagittal localizer. Three dimensional (3D) high resolution T1-weighted images were acquired for anatomical localization (repetition time (TR)=1970 ms, echo time (TE)=4.38 s, flip angle=15°, voxel size=1 × 1 × 1 mm; matrix=256 × 256). A set of two dimensional T1-weighted inversion time echo-planar images consisting of 25 contiguous slices was acquired before each functional run (TR=600 ms; TE=15 ms; flip angle=90°, matrix=128 × 256; in-plane resolution of 1.5 × 0.8 × 4 mm with 1 mm gap). For functional scans, T2*-weighted echo-planar image with BOLD contrast (Kwong et al., 1992; Ogawa *et al*., 1992) were acquired (TR=2500 ms, TE=60 ms, flip angle=80°; matrix =64 × 64; voxel size=3 × 3 × 4 mm with 1 mm gap). A total of 108 volumes were acquired for each functional run.

### 2.6 Data analysis

Baseline demographic and clinical data were compared between groups using independent t-tests or χ2 analyses. fMRI data were analyzed using the FSL software (FMRIB Software Library 3.1). Image preprocessing steps included: (1) spatial realignment to the center volume for motion correction, (2) co-registration of functional images with the high-resolution structural scan using a seven-parameter rigid body transformation, and (3) spatial smoothing using a five-mm full width-half-maximum Gaussian kernel. Two a *priori* ROIs associated with neuroplastic changes after stroke [bilateral MI and PMd cortices] were selected. MI is defined as the gyrus between central sulcus and precentral sulcus including the hand knob. PMd includes the gyrus dorsal from the precentral sulcus not exceeding 10mm. Cluster-based activation Z-maps were constructed to calculate the mean number of activated voxels with a threshold of Z>3.1 (corresponding to a P-value <0.01, corrected for multiple comparisons) in both ROIs.

Voxel counts were computed and used to calculate a laterality index for each ROI [LI = (C-I)/C+I)]; where C and I indicate contralateral (lesioned) and ipsilateral (non-lesioned) activation to the grasping hand, respectively. LI ranges from 1 (activation only of the ROI of the lesioned hemisphere) to -1 (activation only of the ROI of the non-lesioned hemisphere). LI change scores (post-pre) were calculated for each ROI, in each group, for each grasp condition. If the voxel count number for both hemispheres were zero for a given participant, LI was not calculated for that condition.

### 2.7 Statistical analysis

Analyses were conducted using the R statistical computing package (version 3.5.1). Within-group behavioral data were compared using paired t-tests. A multiple linear regression was performed to estimate the task-specific effects of the intervention on ΔLI in two separate models (PMd and MI). To test the hypothesis that the precision task, more than the power, would elicit greater activation of the lesioned PMd/MI in the CIMT compared to the non-CIMT control group, we included an interaction term (group × task) and performed post-hoc t-test comparison to determine the locus of the interaction. Standard errors and 95% confidence intervals (CI) for regression estimates were confirmed over 1,000 bootstrap replicates. Using a backward selection approach, we included potential confounding variables—age, sex, chronicity, lesion volume, and FMA—one at a time and preserved any variable that met a liberal cut-off of p=0.2. Continuous variables were assessed for normality using QQ plots and Shapiro-Wilk tests. Of these, the distribution for lesion volume was extremely positively skewed and was log-transformed. None of the potential confounding variables met the significance cut-off criterion and were therefore not included in the final model. All necessary assumptions for generalized linear models were tested when appropriate.

## 3. Results

No statistically significant differences were found between groups for any demographic or stroke characteristics at baseline (Table 1), except for the MAL QOM which was lower for the non-CIMT group. LI was not computed for two participants in the non-CIMT group (one for the precision task only and one for both tasks) due to a voxel count of zero in at least one hemisphere. Lesion size and location varied in both groups (Table S1). Limb concordance was reported in 3/8 CIMT and 2/6 non-CIMT participants.

**Table 1.**
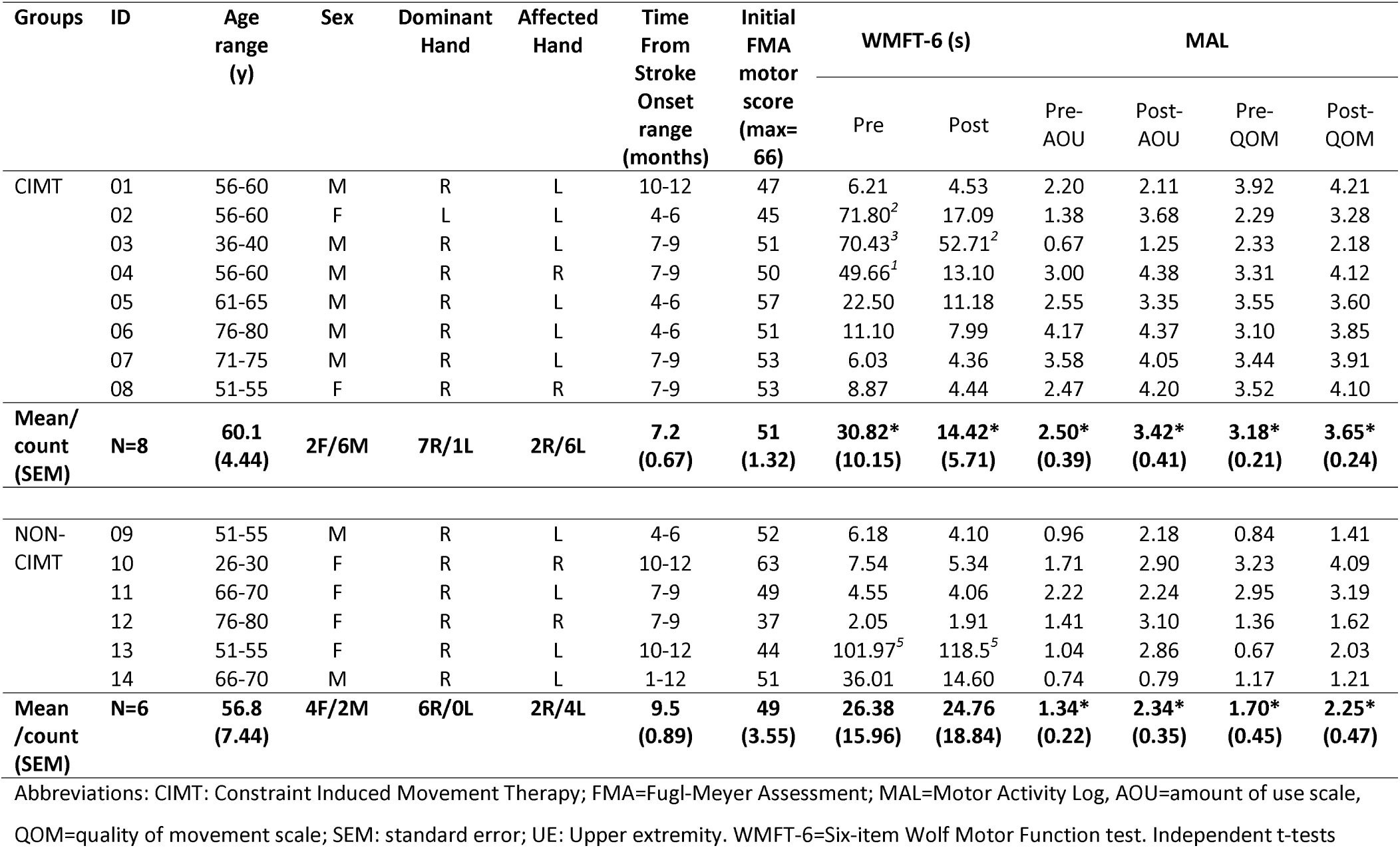
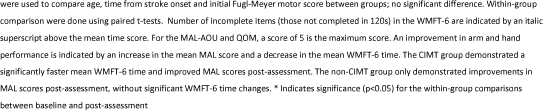
Characteristics of the constraint-induced movement therapy (CIMT) group and non-CIMT group and effect of CIMT treatment on behavioral outcomes

| Groups | ID | Age range (y) | Sex | Dominant Hand | Affected Hand | Time From Stroke Onset range (months) | Initial FMA motor score (max= 66) | WMFT-6 (s) |  | MAL |  |  |  |
| --- | --- | --- | --- | --- | --- | --- | --- | --- | --- | --- | --- | --- | --- |
|  |  |  |  |  |  |  |  | Pre | Post | Pre-AOU | Post-AOU | Pre-QOM | Post-QOM |
| CIMT | 01 | 56-60 | M | R | L | 10-12 | 47 | 6.21 | 4.53 | 2.20 | 2.11 | 3.92 | 4.21 |
|  | 02 | 56-60 | F | L | L | 4-6 | 45 | 71.80<sup>2</sup> | 17.09 | 1.38 | 3.68 | 2.29 | 3.28 |
|  | 03 | 36-40 | M | R | L | 7-9 | 51 | 70.43<sup>3</sup> | 52.71<sup>2</sup> | 0.67 | 1.25 | 2.33 | 2.18 |
|  | 04 | 56-60 | M | R | R | 7-9 | 50 | 49.66<sup>1</sup> | 13.10 | 3.00 | 4.38 | 3.31 | 4.12 |
|  | 05 | 61-65 | M | R | L | 4-6 | 57 | 22.50 | 11.18 | 2.55 | 3.35 | 3.55 | 3.60 |
|  | 06 | 76-80 | M | R | L | 4-6 | 51 | 11.10 | 7.99 | 4.17 | 4.37 | 3.10 | 3.85 |
|  | 07 | 71-75 | M | R | L | 7-9 | 53 | 6.03 | 4.36 | 3.58 | 4.05 | 3.44 | 3.91 |
|  | 08 | 51-55 | F | R | R | 7-9 | 53 | 8.87 | 4.44 | 2.47 | 4.20 | 3.52 | 4.10 |
| Mean/<br>count<br>(SEM) | N=8 | 60.1<br>(4.44) | 2F/6M | 7R/1L | 2R/6L | 7.2<br>(0.67) | 51<br>(1.32) | 30.82*<br>(10.15) | 14.42*<br>(5.71) | 2.50*<br>(0.39) | 3.42*<br>(0.41) | 3.18*<br>(0.21) | 3.65*<br>(0.24) |
| NON-CIMT | 09 | 51-55 | M | R | L | 4-6 | 52 | 6.18 | 4.10 | 0.96 | 2.18 | 0.84 | 1.41 |
|  | 10 | 26-30 | F | R | R | 10-12 | 63 | 7.54 | 5.34 | 1.71 | 2.90 | 3.23 | 4.09 |
|  | 11 | 66-70 | F | R | L | 7-9 | 49 | 4.55 | 4.06 | 2.22 | 2.24 | 2.95 | 3.19 |
|  | 12 | 76-80 | F | R | R | 7-9 | 37 | 2.05 | 1.91 | 1.41 | 3.10 | 1.36 | 1.62 |
|  | 13 | 51-55 | F | R | L | 10-12 | 44 | 101.97<sup>5</sup> | 118.5<sup>5</sup> | 1.04 | 2.86 | 0.67 | 2.03 |
|  | 14 | 66-70 | M | R | L | 1-12 | 51 | 36.01 | 14.60 | 0.74 | 0.79 | 1.17 | 1.21 |
| Mean<br>/count<br>(SEM) | N=6 | 56.8<br>(7.44) | 4F/2M | 6R/0L | 2R/4L | 9.5<br>(0.89) | 49<br>(3.55) | 26.38<br>(15.96) | 24.76<br>(18.84) | 1.34*<br>(0.22) | 2.34*<br>(0.35) | 1.70*<br>(0.45) | 2.25*<br>(0.47) |

Abbreviations: CIMT: Constraint Induced Movement Therapy; FMA=Fugl-Meyer Assessment; MAL=Motor Activity Log, AOU=amount of use scale, QOM=quality of movement scale; SEM: standard error; UE: Upper extremity. WMFT-6=Six-item Wolf Motor Function test. Independent t-tests
were used to compare age, time from stroke onset and initial Fugl-Meyer motor score between groups; no significant difference. Within-group comparison were done using paired t-tests. Number of incomplete items (those not completed in 120s) in the WMFT-6 are indicated by an italic superscript above the mean time score. For the MAL-AOU and QOM, a score of 5 is the maximum score. An improvement in arm and hand performance is indicated by an increase in the mean MAL score and a decrease in the mean WMFT-6 time. The CIMT group demonstrated a significantly faster mean WMFT-6 time and improved MAL scores post-assessment. The non-CIMT group only demonstrated improvements in MAL scores post-assessment, without significant WMFT-6 time changes. \* Indicates significance ( $p < 0.05$ ) for the within-group comparisons between baseline and post-assessment

At the behavioral level, participants in the CIMT group showed significant improvements in both the WMFT-6 (t= 2.382, p=0.049) and the MAL post intervention (AOU: t=-3.199, p=0.015; QOM: t=-3.482, p=0.011). The control group showed only improvements in the MAL (AOU: t=-3.108, p=0.027; QOM: t=2.783, p=0.039).

### 3.1 Precision task elicits greater relative activation of PMd of the lesioned hemisphere in CIMT group compared to controls

Our final model for the PMd ROI was significantly different from a null model (F(4,20)=4.65, p=0.012, adj. R^2^ =32.3%). However, that for the MI ROI was not significant (F(4,20)=1.05, p =0.389, adj. R ^2^=0.72%).

Consistent with our hypothesis, there was a significant interaction between group and task (B=1.31, p=0.004). The post-hoc analysis confirmed the locus of the interaction; specifically for the precision task, the CIMT group showed an increase PMd ΔLI, i.e., increased activation of the PMd of the lesioned hemisphere relative to the non-lesioned hemisphere, compared to the non-CIMT group (t=3.458, p=0.002; Figure 2A). This group level change was apparent on an individual level (Figure 2B); PMd ΔLI increased towards the lesioned hemisphere in 6/7 participants for the CIMT group, while LI decreased in 5/6 participants for the non-CIMT group.

**Figure 2.**
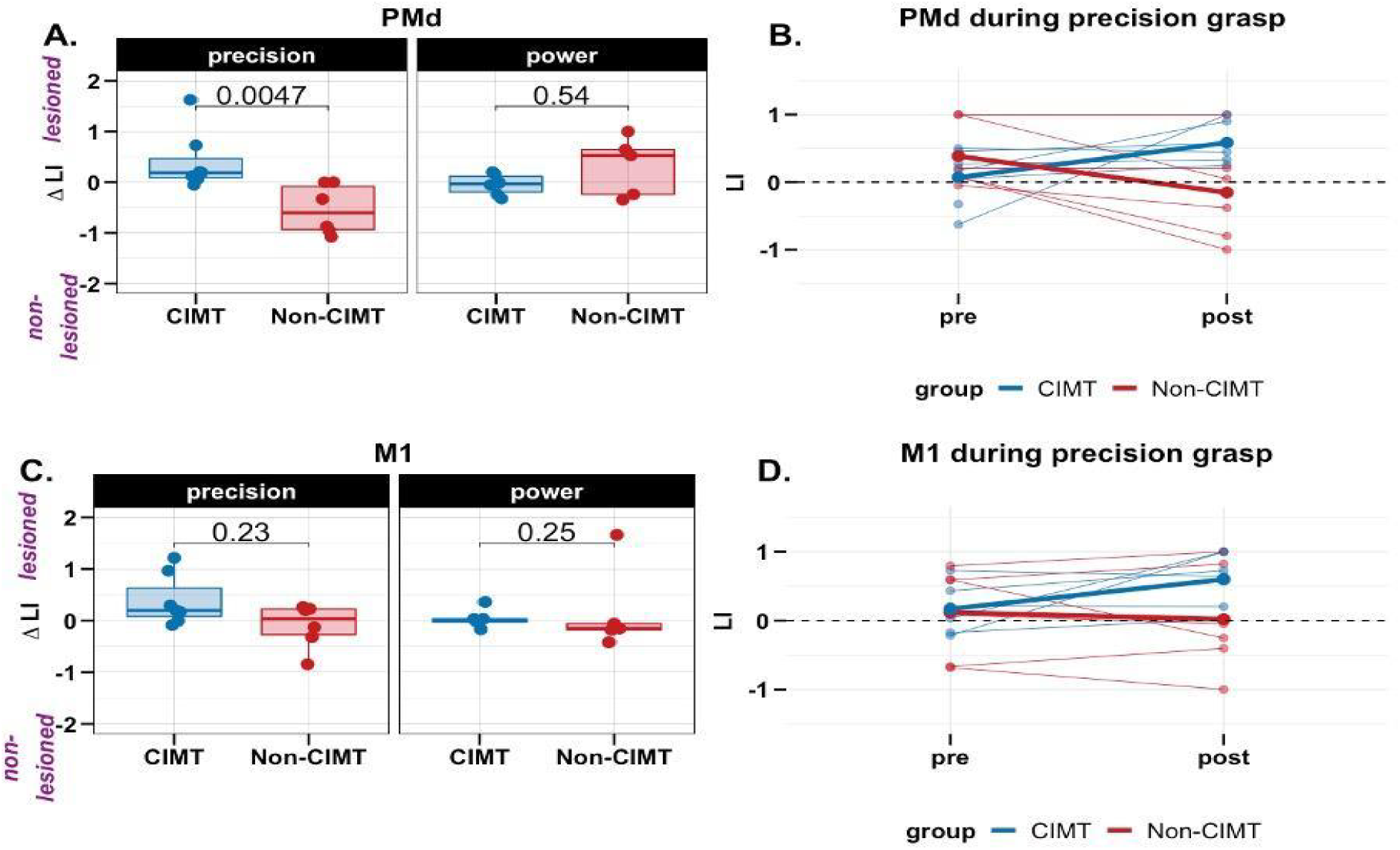
Change in laterality index (LI) across groups and tasks (positive values in ΔLI indicate greater activation of the ROI on the lesioned relative to non-lesioned hemisphere). A) For the precision task, compared to the non-CIMT group (n = 6), the constraint-induced movement therapy (CIMT) group (n = 7) showed increased activation of the dorsal premotor cortex (PMd) in the lesioned hemisphere. However, this effect was not observed for the power task (CIMT group, n = 6, non-CIMT group, n = 5). B) Individual changes in LI from pre-post in PMd; thicker lines are group means C) In the primary motor cortex (MI), no changes in LI were observed from pre to post for both groups; D) Individual changes in LI from pre-post in MI; thicker lines are group means.

There was also a significant main effect of ‘*task*’ (Table 2) such that compared to the power task, PMd ΔLI was smaller for the precision task, when averaged between the two groups. Change in activation of PMd tended to be smaller for the precision compared to the power grasp. However, given the strong interaction between group and task (i.e., the strong effect of group on PMd ΔLI for the precision but not power task), interpreting this main effect on its own, by averaging between groups, is misleading.

**Table 2.** Estimates from multiple linear regression.

| Predictors | PMd ΔLI | | | MI ΔLI | | |
| --- | --- | --- | --- | --- | --- | --- |
|  | Estimates | CI | p | Estimates | CI | p |
| (Intercept) | 0.32 | -0.14-0.78 | 0.166 | 0.17 | -0.32-0.66 | 0.473 |
| Group | -0.36 | -0.98-0.26 | 0.241 | -0.13 | -0.79-0.52 | 0.675 |
| Task | -0.86 | -1.48- -0.23 | **0.010** | -0.27 | -0.93-0.39 | 0.402 |
| Group x Task | 1.31 | 0.46-2.16 | **0.004** | 0.63 | -0.27-1.52 | 0.159 |
| Observations | 24 |  |  | 24 |  |  |
| R<sup>2</sup>/ Adjusted R<sup>2</sup> | 0.411/0.323 |  |  | 0.137/0.007 |  |  |

As noted earlier, for the MI ROI, there was no significant difference between CIMT and non-CIMT for either task (Figures 2C & D).

## 4. Discussion

In this retrospective analysis, we found that compared to a no-treatment control group, two weeks of CIMT resulted in a relative increase in activity in a key node of the motor network, the PMd of the lesioned hemisphere under precision grasp task conditions. The results underscore the importance of task-specific training in the context of CIMT and its potential for driving motor network activity towards restoration by increasing paretic arm function and hand use, as supported by significant improvements in WMFT-6 and both scales of the MAL. However, contrary to our hypothesis, no significant changes in MI brain activation were observed for the precision or power tasks in either group. These findings are preliminary in view of the small sample size, stressing the need to be replicated in a larger and similarly rigorous study.

The PMd ΔLI findings corroborates the effect of CIMT in driving neuroplasticity. These results were complimented by behavioral changes both in precision task performance (WMFT-6) and perceived quantity and quality of arm and hand activities (MAL) in the CIMT group. The MAL improvement for the non-CIMT group was likely induced through initial exposure to the MAL battery of questions, drawing participants’ attention to the paretic side and its use, since there was no real change in the WMFT-6 task performance measures. Previous studies report evidence for cortical reorganization in sensorimotor areas induced by task-specific training (Levy *et al*., 2001; Johansen-Berg et al., 2002; Jang *et al*., 2003; Kim *et al*., 2004), but do not differentiate between key neural nodes within the sensorimotor network associated with specific task conditions. Our results are consistent with previous work that emphasizes the role of PMd in recovery mechanisms and the precise regulation of force (Johansen-Berg *et al*., 2002; Ward *et al*., 2007; Bestmann *et al*., 2010). Consistent with the results from Bestmann *et al*. (2010), input from lesioned hemisphere PMd might assist the lesioned brain regions to produce movement. Evidence supports the role of PMd in movement execution, especially for stroke survivors with greater motor impairments (Ward *et al*., 2007), and in the control of skilled movement beyond simple execution, that which involves movement planning (Kantak *et al*., 2012; Stewart *et al*., 2016). One interpretation is that CIMT because of its repetitive and progressively more challenging practice, elicits motor learning and decision-making processes, including anticipatory action planning, which uniquely engages PMd in the selection of action. In our previous work (Tan *et al*., 2012), we provided evidence that the CIMT group demonstrated more optimal anticipatory hand posturing prior to precision grasp tasks than did the control group. These findings combined with our behavioral results here, complement the current fMRI findings and support the differential effect of CIMT on PMd. Our results also provide partial support for the hypothesis that the recovery of hand motor function following a stroke is mediated by separate systems for strength and dexterity (Xu *et al*., 2017; Mawase *et al*., 2019).

One aspect of our PMd laterality index change findings that surprised us was the apparent *decrease* in LI, toward the non-lesioned hemisphere that we observed for the no-treatment control group (red line in Figure 2B). One possible interpretation based on our behavioral results and mentioned earlier is that the mere exposure to the MAL questionnaire at baseline primes the individual to attempt more use of the paretic limb in everyday activities, especially for bimanual tasks as a support or stabilizer, but not necessarily for dexterous activities that would be emphasized as part of CIMT. If this was the case, that increased use could have manifested as compensatory activation of the non-lesioned hemisphere PMd and as increased perceived use and quality of use of the paretic limb that we noted for MAL amount and quality scores (Uswatte *et al*., 2006; Narai *et al*., 2016) (Table 1). The fact that we did not see significant control group improvements for dexterous performance of the WMFT-6 is consistent with this explanation.

This study adds to the literature by demonstrating that cortical reorganization associated with CIMT is a function of the specific tasks practiced and can be restorative rather than compensatory in nature (Hodics, Cohen and Cramer, 2006; Levin, Kleim and Wolf, 2009). The novelty of our study is that neural plastic changes were examined as a function of task type, unlike most studies that have focused primarily on whole hand grasp tasks.

Unlike PMd, we did not observe significant MI ΔLI for either group or task. We suggest the following interpretation for these null findings. The force level (i.e. pressure) was kept constant over repeated fMRI sessions to control for the well-known relationship between force (i.e., muscle torque) and MI activation, and for performance (i.e. strength) gains that were expected to occur in the CIMT group but not necessarily in the non-CIMT group (Cramer *et al*., 2002). Our careful control to obtain consistent force and rate levels between scans may have biased the results towards observing differences in motor planning rather than execution.

Another potential confounding factor is that two participants, one in the CIMT group and one in the non-CIMT group, had brain lesions directly affecting MI, but none had lesions directly affecting PMd. The task-specific nature of CIMT training may induce important changes upstream from M1 in motor areas responsible for higher-level task planning, including movement preparation and action selection, those functions for which PMd has an important role (Grafton, Fagg and Arbib, 1998; Hoshi and Tanji, 2007; Kantak *et al*., 2012). Evidence supports that the strengthened functional connectivity between PMd and MI in the ipsilesional hemisphere is associated with improved long-term retention of motor skills (Lefebvre *et al*., 2015, 2017). Taken together, the choice of MI for one of two pre-specified ROIs was perhaps naïve given that CIMT does not directly target hand strength and importantly, we constrained the force and rate levels to maintain consistent performance across fMRI sessions.

No study is without limitations. The fMRI data were collected using a 1.5 Tesla scanner, which produced lower image quality than what is now used in research. Given the high variability in the lesion locations and clinical presentation of stroke, the main limitation of this study is its small sample size. The lack of sufficient samples combined with the large variability across participants limits generalizability of our findings. Future studies should examine the relationship between the effects demonstrated here and lesion size and location. It is possible that performance variability across participants during imaging added noise to the data that overwhelmed the signal. However, implementation of a rigorous pre-training phase outside of the scanner and individual-participant task performance criteria (% max force and rate) promoted performance consistency across scanning sessions and reduced the likelihood that activation was due to performance variability. On the other hand, this rigorous pre-training may explain the null effects observed for MI compared with those for PMd (i.e., differences in motor planning rather than execution).

Intense task-specific training of the affected limb in combination with restraint of the less-affected limb drives functional reorganization by shifting bi-hemispheric motor cortical activity toward the lesioned hemisphere. These results provide strong evidence from humans undergoing rehabilitation for the principle of specificity and intensity of experience-dependent plasticity derived primarily from animal models in the research laboratory (Kleim and Jones, 2008). These findings should be replicated in a larger study.

## Data Availability

The complete codebook for analysis is available through the first author's OSF repository: https://osf.io/89mkd/

https://osf.io/89mkd/

## 5. Conflict of Interest

The authors declare that the research was conducted in the absence of any commercial or financial relationships that could be construed as a potential conflict of interest.

## 6. Author Contributions

**Marika Demers:** Writing- Original draft preparation and visualization.

**Rini Varghese:** Formal analysis, software, data curation, visualization and writing-reviewing and editing.

**Carolee Winstein:** conceptualization, supervision, writing-reviewing and editing.

## 7. Funding

The study was supported by National Institutes of Health grants R01 NS 45485 to Dr. Carolee Winstein, R24 HD 39629 (Western Medical Rehabilitation Research Network) to Dr. Bruce Dobkin and F31HD098796 to Dr. Rini Varghese. The study was also supported by the post-doctoral fellowship from the Fonds de la Recherche du Québec Santé (#268272) to Dr. Marika Demers. The study was supported by funding from the Ahmanson-Lovelace Brain Mapping Center, the National Center for Research Resources grants RR12169, RR13642, and RR08655 NIH NIDA K12DA000167 and American Heart Association 14CRP18200010. The funding sources did not have a role in the redaction of this manuscript.

## 8. Acknowledgments

We thank Allan Wu and Bruce Dobkin at the University of California, Los Angeles, and Indu Sitarju and Katie Lerch from the University of Southern California (USC) for their assistance during data collection and aspects of data analysis. We thank the USC clinical team of Michelle Prettyman, Samantha Underwood, Chris Hahn, Janice Lin, and Jarugool Tretriluxana for participant recruitment and behavioral assessments, and Dorsa Beroukhim Kay, Kathleen Garrison, Yun Dong, Matthew Konersman, Steve Cen, Vikas Rao, and Nicolo Betoni for post processing, analysis of the fMRI data, and for brainstorming sessions over the years about these data. We thank Michael Callegari for the custom software used for pneumatic-force calibration and grasp task analysis. A pre-print of this manuscript was published on medRxiv (Demers, Varghese and Winstein, 2021).

## 10. Data Availability Statement

The complete codebook for statistical analysis is available through the first author’s OSF repository: https://osf.io/89mkd/

## Notes

### Competing Interest Statement

The authors have declared no competing interest.

### Clinical Trial

NCT04989920

### Funding Statement

The study was supported by National Institutes of Health grants NS 45485 to Dr. Carolee Winstein, R24 HD 39629 (Western Medical Rehabilitation Research Network) to Dr. Bruce Dobkin and F31HD098796 to Dr. Rini Varghese. The study was also supported by the post-doctoral fellowship from the Fonds de la Recherche du Quebec Sante (#268272) to Dr. Marika Demers. The study was supported by funding from the Ahmanson-Lovelace Brain Mapping Center, the National Center for Research Resources grants RR12169, RR13642, and RR08655 NIH NIDA K12DA000167 and American Heart Association 14CRP18200010. The funding sources did not have a role in the redaction of this manuscript.

### Author Declarations

University of Southern California (USC) Health Sciences Campus IRB, HS-016022.

### Summary of Updates

Data collection and results updated to include behavioral data.

## References

Bestmann, S. et al. (2010) ‘The Role of Contralesional Dorsal Premotor Cortex after Stroke as Studied with Concurrent TMS-fMRI’, Journal of Neuroscience, 30(36), pp. 11926–11937. doi:10.1523/JNEUROSCI.5642-09.2010.

Biernaskie, J. and Corbett, D. (2001) ‘Enriched rehabilitative training promotes improved forelimb motor function and enhanced dendritic growth after focal ischemic injury’, The Journal of neuroscience: the official journal of the Society for Neuroscience, 21(14), pp. 5272–5280. doi:10.1523/JNEUROSCI.21-14-05272.2001.

Buma, F.E. et al. (2010) ‘Review: Functional Neuroimaging Studies of Early Upper Limb Recovery After Stroke: A Systematic Review of the Literature’, Neurorehabilitation and Neural Repair, 24(7), pp. 589–608. doi:10.1177/1545968310364058.

Calautti, C. et al. (2010) ‘The relationship between motor deficit and primary motor cortex hemispheric activation balance after stroke: longitudinal fMRI study’, Journal of Neurology, Neurosurgery & Psychiatry, 81(7), pp. 788–792. doi:10.1136/jnnp.2009.190512.

Carey, J.R., Bhatt, E. and Nagpal, A. (2005) ‘Neuroplasticity promoted by task complexity’, Exercise and Sport Sciences Reviews, 31, pp. 24–31.

Castro-Alamancos, M.A. and Borrell, J. (1995) ‘Functional recovery of forelimb response capacity after forelimb primary motor cortex damage in the rat is due to the reorganization of adjacent areas of cortex’, Neuroscience, 68(3), pp. 793–805. doi:10.1016/0306-4522(95)00178-L.

Corbetta, D. et al. (2015) ‘Constraint-induced movement therapy for upper extremities in people with stroke’, Cochrane Database of Systematic Reviews, 10. doi:10.1002/14651858.CD004433.pub3.

Cramer, S.C. et al. (2002) ‘Motor cortex activation is related to force of squeezing’, Human Brain Mapping, 16(4), pp. 197–205. doi:10.1002/hbm.10040.

Demers, M., Varghese, R. and Winstein, C.J. (2021) ‘Retrospective Exploratory Analysis of Task-Specific Effects on Brain Activity after Stroke’, MedRxiv [Preprint]. doi:10.1101/2021.08.20.21260371.

Fugl-Meyer, A.R. et al. (1975) ‘The post stroke hemiplegic patient. I. A method for evaluation of physical performance’, Scandinavian Journal of Rehabilitation Medicine, 7(1), pp. 13–31.

Grafton, S.T., Fagg, A.H. and Arbib, M.A. (1998) ‘Dorsal Premotor Cortex and Conditional Movement Selection: A PET Functional Mapping Study’, Journal of Neurophysiology, 79(2), pp. 1092–1097.

Hodics, T., Cohen, L. and Cramer, S. (2006) ‘Functional Imaging of Intervention Effects in Stroke Motor Rehabilitation’, Archives of Physical Medicine and Rehabilitation, 87(12), pp. 36–42. doi:10.1016/j.apmr.2006.09.005.

Hoshi, E. and Tanji, J. (2007) ‘Distinctions between dorsal and ventral premotor areas: anatomical connectivity and functional properties’, Current Opinion in Neurobiology, 17(2), pp. 234–242. doi:10.1016/j.conb.2007.02.003.

Jang, S.H. et al. (2003) ‘Cortical reorganization induced by task-oriented training in chronic hemiplegic stroke patients’, Neuroreport, 14(1), pp. 137–141. doi:10.1097/01.wnr.0000051544.96524.f2.

Johansen-Berg, H. et al. (2002) ‘The role of ipsilateral premotor cortex in hand movement after stroke’, Proceedings of the National Academy of Sciences, 99(22), pp. 14518–14523. doi:10.1073/pnas.222536799.

Jones, T.A. et al. (1999) ‘Motor skills training enhances lesion-induced structural plasticity in the motor cortex of adult rats’, The Journal of Neuroscience, 19(22), pp. 10153–10163.

Kantak, S.S. et al. (2012) ‘Rewiring the brain: Potential role of the premotor cortex in motor control, learning, and recovery of function following brain injury’, Neurorehabilitation and Neural Repair, 26(3), pp. 282–292. doi:10.1177/1545968311420845.

Kaplon, R. et al. (2007) ‘Six hours in the laboratory? A quantification of practice time during Constraint-Induced Therapy’, Clinical Rehabilitation, 21, pp. 950–958. doi:10.1177/0269215507078333.

Kim, Y.H. et al. (2004) ‘Plastic changes of motor network after constraint-induced movement therapy’, Yonsei Medical Journal, 45(2), pp. 241–246. doi:200404241 [pii].

Kleim, J.A. and Jones, T.A. (2008) ‘Principles of experience-dependent neural plasticity: implications for rehabilitation after brain damage’, Journal of Speech, Language, and Hearing Research, 51(1), pp. S225–S239.

Kwakkel, G. et al. (2015) ‘Constraint-induced movement therapy after stroke’, The Lancet Neurology, 14(2), pp. 224–234. doi:10.1016/S1474-4422(14)70160-7.

Kwong, K.K. et al. (1992) ‘Dynamic magnetic resonance imaging of human brain activity during primary sensory stimulation.’, Proceedings of the National Academy of Sciences of the United States of America, 89(12), pp. 5675–5679. doi:10.1073/pnas.89.12.5675.

Laible, M. et al. (2012) ‘Association of activity changes in the primary sensory cortex with successful motor rehabilitation of the hand following stroke’, Neurorehabilitation and Neural Repair, 26(7), pp. 881–888. doi:10.1177/1545968312437939.

Lefebvre, S. et al. (2015) ‘Neural substrates underlying stimulation-enhanced motor skill learning after stroke’, Brain, 138(1), pp. 149–163. doi:10.1093/brain/awu336.

Lefebvre, S. et al. (2017) ‘Increased functional connectivity one week after motor learning and tDCS in stroke patients’, Neuroscience, 340, pp. 424–435. doi:10.1016/j.neuroscience.2016.10.066.

Levin, M.F., Kleim, J.A. and Wolf, S.L. (2009) ‘What do motor “recovery” and “compensation” mean in patients following stroke?’, Neurorehabilitation Neural Repair. 2009/01/02 edn, 23(4), pp. 313–319. doi:1545968308328727 [pii] 10.1177/1545968308328727.

Levy, C.E. et al. (2001) ‘Functional MRI evidence of cortical reorganization in upper-limb stroke hemiplegia treated with constraint-induced movement therapy’, American Journal of Physical Medicine and Rehabilitation, 80(1), pp. 4–12.

Liepert, J. et al. (2006) ‘The surround inhibition determines therapy-induced cortical reorganization’, NeuroImage, 32(3), pp. 1216–1220. doi:10.1016/j.neuroimage.2006.05.028.

Mawase, F. et al. (2019) ‘Corticocortical systems underlying high-order motor control’, Brain, 91(4), pp. 1722–1733. doi:10.1152/jn.00805.2003.

Muir, R.B. and Lemon, R.N. (1983) ‘Corticospinal neurons with a special role in precision grip’, Brain Research, 261(2), pp. 312–316. doi:10.1016/0006-8993(83)90635-2.

Napier, J.R. (1956) ‘The prehensive movements of the human hand’, The Journal of Bone and Joint Surgery, 38, pp. 902–913. doi:10.1302/0301-620X.38B4.902.

Narai, E. et al. (2016) ‘Accelerometer-based monitoring of upper limb movement in older adults with acute and subacute Stroke’, Journal of Geriatric Physical Therapy, 39(4), pp. 171–177. doi:10.1519/JPT.0000000000000067.

Nudo, R.J. et al. (1996) ‘Neural substrates for the effects of rehabilitative training on motor recovery after ischemic infarct’, Science, 272(5269), pp. 1791–1794. doi:10.1126/science.272.5269.1791.

Nudo, R.J. (2007) ‘Postinfarct Cortical Plasticity and Behavioral Recovery’, Stroke, 38(2), pp. 840–845. doi:10.1161/01.STR.0000247943.12887.d2.

Ogawa, S. et al. (1992) ‘Intrinsic signal changes accompanying sensory stimulation: functional brain mapping with magnetic resonance imaging.’, Proceedings of the National Academy of Sciences of the United States of America, 89(13), pp. 5951–5955. doi:10.1073/pnas.89.13.5951.

Schaechter, J.D. et al. (2002) ‘Motor recovery and cortical reorganization after constraint-induced movement therapy in stroke patients: a preliminary study’, Neurorehabilitation and Neural Repair, 16(4), pp. 326–338. doi:10.1177/154596830201600403.

Schaechter, J.D. and Perdue, K.L. (2008) ‘Enhanced cortical activation in the contralesional hemisphere of chronic stroke patients in response to motor skill challenge’, Cerebral Cortex, 18(3), pp. 638–647. doi:10.1093/cercor/bhm096.

Stewart, J.C. et al. (2016) ‘Dorsal premotor activity and connectivity relate to action selection performance after stroke’, Human Brain Mapping, 37(5), pp. 1816–1830. doi:10.1002/hbm.23138.

Tan, C. et al. (2012) ‘Anticipatory planning of functional reach-to-grasp: A pilot study’, Neurorehabilitation and Neural Repair, 26(8), pp. 957–967. doi:10.1177/1545968312437938.

Taub, E. et al. (1993) ‘Technique to improve chronic motor deficit after stroke’, Archives of Physical Medicine and Rehabilitation, 74(4), pp. 347–354.

Taub, E. et al. (2013) ‘Method for enhancing real-world use of a more affected arm in chronic stroke’, Stroke, 44(5), pp. 1383–1388. doi:10.1161/STROKEAHA.111.000559.

Uswatte, G. et al. (2006) ‘Validity of accelerometry for monitoring real-world arm activity in patients with subacute stroke: evidence from the Extremity Constraint-Induced Therapy Evaluation Trial’, Archives of Physical Medicine and Rehabilitation, 87(10), pp. 1340–1345. doi:10.1016/j.apmr.2006.06.006.

Uswatte, G. and Taub, E. (1999) ‘Constraint-induced movement therapy: New approaches to outcome measurement in rehabilitation’, in Cognitive neurorehabilitation. New York, NY, US: Cambridge University Press, pp. 215–229.

Ward, N.S. et al. (2007) ‘The relationship between brain activity and peak grip force is modulated by corticospinal system integrity after subcortical stroke’, European Journal of Neuroscience, 25(6), pp. 1865–1873. doi:10.1111/j.1460-9568.2007.05434.x.

Winstein, C.J. et al. (2003) ‘Methods for a Multisite Randomized Trial to Investigate the Effect of Constraint-Induced Movement Therapy in Improving Upper Extremity Function among Adults Recovering from a Cerebrovascular Stroke’, Neurorehabilitation and Neural Repair, 17(3), pp. 137–152. doi:10.1177/0888439003255511.

Winstein, C.J. and Wolf, S.L. (2014) ‘Task-oriented training to promote upper extremity recovery’, in Stein J Macko RF, Winstein CJ, Zorowitz RD H.R.L,. (ed.) Stroke Recovery and Rehabilitation. 2nd edition. New York: Demos Medical, pp. 267–290.

Wittenberg, G.F. et al. (2003) ‘Constraint-Induced Therapy in Stroke: Magnetic-Stimulation Motor Maps and Cerebral Activation’, Neurorehabilitation and Neural Repair, 17(1), pp. 48–57. doi:10.1177/0888439002250456.

Wolf, S.L. et al. (2005) ‘The EXCITE trial: Attributes of the Wolf Motor Function Test in patients with subacute stroke’, Neurorehabilitation and Neural Repair, 19(3), pp. 194–205. doi:10.1177/1545968305276663.

Wolf, S.L. et al. (2008) ‘The EXCITE Trial: Retention of Improved Upper Extremity Function Among Stroke Survivors Receiving CI Movement Therapy’, Lancet neurology, 7(1), pp. 33–40. doi:10.1016/S1474-4422(07)70294-6.

Xu, J. et al. (2017) ‘Separable systems for recovery of finger strength and control after stroke’, Journal of Neurophysiology, 118(2), pp. 1151–1163. doi:10.1152/jn.00123.2017.

