## Supplemental Table S1 for "Retrospective Exploratory Analysis of Task-Specific Effects on Brain Activity after Stroke" (from another version of this preprint: version 1, 2021-08-25)

**Supplementary Table S1:** Lesion description

| | ID | 3D Image | 3D Volume (cc) Location | ID | 3D Image | 3D Volume (cc) Location |
|---|---|---|---|---|---|---|
| CIMT | 01 | 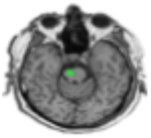 | 0.2 R Pontine | 05 | 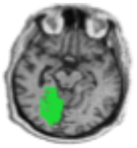 | 14.8 R O |
| | 02 | 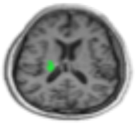 | 0.7 R IC | 06 | 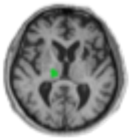 | 1.2 R IC |
| | 03 | 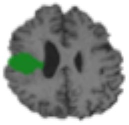 | 14.5 R M1 | 07 | 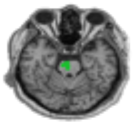 | 0.8 R Pontine |
| | 04 | 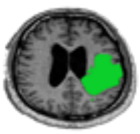 | 72.6 R P,T | 08 | 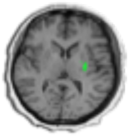 | 0.9 L IC |
| Non-CIMT | 09 | 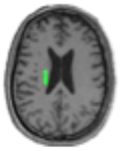 | 0.7 R CN,CR | 12 | 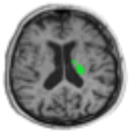 | 1.4 L CN,CR |
| | 10 | 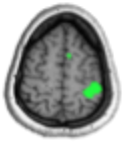 | 3.1 L M1 | 13 | 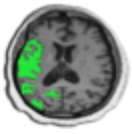 | 27.7 R SM,T |
| | 11 | 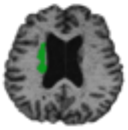 | 2.3 2R CN, CR | 14 | 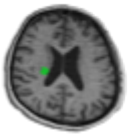 | 0.8 R CR,CN |

**Legend:** CIMT, constraint-induced movement therapy; R, Right; L, Left; B, brainstem; F, Frontal area; T, Temporal area; P, Parietal area; O, Occipital area; M1, primary motor area; SM, Sensorimotor area; CR, Corona radiata; CN, Caudate nucleus; IC, Internal capsule. *The green area in the 3D image shows the lesion on the slice with the largest computed area.
